# Does a non-targeted publicly-funded health care voucher system for the elderly improve access to optometry services?

**DOI:** 10.1101/2020.04.01.20049445

**Authors:** Rita Sum, Maurice Yap

## Abstract

**Objective:** To study how a non-targeted publicly-funded health care voucher system for the elderly impact on access to optometry services from the perspective of service users and service providers

**Design:** Cross-sectional study

**Setting:** 19 elderly community centers

**Participants:** 1176 people, aged 65 years or above, and 389 optometrists

**Primary and secondary outcome measures:** Usage characteristics of optometry services by eligible service users of the voucher scheme

Perspectives of eligible service users on access barriers to optometry services

Perspectives of service providers on voucher scheme

**Results:** In total, 1156 valid questionnaires were collected from a cohort of eligible service users. Results showed that 53.7% of subjects had used optometry services within the past 2 years, while 22% had not used optometry services before. Lack of familiarity with services provided, professional fees and prices of prescription spectacles were the main barriers to using optometry services. Of those subjects who had used the voucher for optometry service before, 80.4% had eye examination in the past 2 years, versus 64.1% among subjects who had not use health care voucher on the optometry service. “Insufficient voucher value” was a commonly quoted reason for not using the health care vouchers for optometry services. Over 80% of optometrists agreed that the voucher scheme improved the awareness of major eye conditions and enabled the elderly to have prescription spectacles when necessary.

**Conclusion:** The health care voucher for the elderly improved access to optometry services. Access could be improved further by promoting awareness optometry services, location of service providers, price transparency of professional services and prescription spectacles. Responses from optometry services providers are supportive of the view that the voucher scheme improved access to and utilization of preventive care services.

**Article Summary:** *Strengths and limitations of this study:*

- High representativeness of community dwelling older population in Hong Kong.
- The mixed method approach provided a more in-depth investigation of the population.
- One limitation is the generalizability of the results with older people who are members in the community centers.

## INTRODUCTION

Hong Kong operates a dual-track health care system. There is universal health care coverage for all residents which is publicly-funded, supplemented by a thriving private sector for those residents who choose to use private clinics and hospitals. Users of the private sector typically have medical insurance, purchased privately or through employer-provided medical benefits, or pay out of pocket.[1] Primary care medical services are mainly delivered in the private sector (70%) whereas secondary care services are provided in the public and private sectors. Older people comprise the majority of users of the public clinics and hospitals.[2] The ageing population is expected to further challenge the heavily burdened public health care sector.

Visual impairment is an important determinant of the quality of life in older people.[3] The prevalence of visual impairment increases with age.[4] Older people have a higher risk of eye anomalies, including uncorrected refractive error, cataract and age-related macular degeneration.[5, 6] They are recommended to have an annual eye examination in order to screen out visual and eye problems early.[7] In Hong Kong, about 40% of older people have visual impairment in at least one eye and the prevalence is over 70% for those aged 80 or above.[8] The waiting time for a medical eye consultation ranges from 63 to 157 weeks in the various public hospitals.[9] Many of those waiting for an appointment have refractive error.[10] Clearly, there is a need to improve access to vision care services, such as by making better use of the available capacity in the private sector. Financial incentive, in the form of a health care voucher, is one strategy to improve utilization of the different health care services available in the private sector.

The voucher system is a form of demand-side financing which enables eligible users, usually an under-privileged group, to have the purchasing power for different services.[11] A voucher programme typically engages private service providers. The programme introduces competition among service providers and promotes improvement in service quality in order to maintain competitiveness.[12] To facilitate the use of private health care services, the Hong Kong Government launched the Elderly Health Care Voucher (EHV) scheme in 2009. In the latest scheme enhancement of 2019, eligible residents aged 65 years or above would receive HK$ 2,000 annually in a designated account. The balance on this account may accumulate up to HK$ 5,000. The Scheme includes services delivered by 10 different health care professionals, including medical practitioners, TCM practitioners, dentists, optometrists, and registered allied health professions. Eligible users can choose to use the voucher on these private services according to their health needs. Some evaluations have been conducted on the progress and outcome of the EHV scheme. The evaluations show that most people used their voucher for curative care; only 4.8% were used for preventive care. The voucher scheme was considered to be a social welfare programme and the non-service specific voucher led to overwhelming use in curative care.[13-16] In the latest review, it was found that the voucher scheme did not reduce utilization of public clinics and hospitals but encouraged dual utilization of public and private healthcare services.[16]

In the Hong Kong context, primary vision care is mostly delivered by optometrists registered under Part 1 of the Optometrists Register. They number about 1,000, around half the optometrists on the Optometrists Register in 2020.[17] and over 90% work in the private sector.[18] Optometry services provided serves two purposes: prescription and supply of optical aids to optimize vision and an eye examination to screen for sight-threatening conditions.

Optometrists with Part 1 registration were added in the EHV scheme in 2012. The inclusion of optometry services was intended to facilitate greater use of preventive care services concerning eye conditions.[19] Apart from vision assessment services, the voucher may also be used for prescription of spectacles if deemed necessary by the attending optometrist. There are increasing numbers of optometrists enrolling in the scheme, from 152 in 2012, 533 in 2016 to 780 in 2019.[20] Over 78% of eligible optometrists are currently enrolled.

With the substantial lowering of the financial barrier to access, we were interested to know what other barriers were preventing people from using optometry services, in particular, as a preventive service. We were also interested to know the views from the supply side, the optometrists who deliver vision care services, on how the EHV impacts on access and what could be improved to deliver the objectives of the scheme.

## METHOD

This study comprised two parts.

The first part investigated the use of the EHV for optometry services. A population-based cross-sectional survey using an assisted self-administered questionnaire was conducted in 19 community centers for the elderly, covering all 18 districts in Hong Kong. Older people, who were community-dwelling and were eligible users of the EHV, were invited to participate in this study. Cluster sampling was adopted according to the geographically-divided districts in Hong Kong. The number of participants in each district required was calculated based on the proportion of older people living in the district. A sample of older people with different pattern of service utilization was invited to have face-to-face interview for better understanding of their views and experience of optometry services. All the interviews were recorded digitally with the permission of the participants. Transcripts of the recordings were later produced.

The second part of the study collected the views of optometrists about the EHV scheme questionnaire. Apart from recruiting optometrists from the professional associations, snowball sampling was also used to enhance recruitment. At the time of recruitment, it was not known whether they were enrolled as service providers in the EHV scheme. The optometrists were asked to complete a cross-sectional survey using a self-administered questionnaire conducted online.

Test-retest reliability of the questionnaires was assessed by inviting 20 participants to complete part of the questionnaire again one week after the first trial. Percentage agreement shows the consistency of the testing results over time.[21] Agreement of 70% or above is considered satisfactory.[22] Bland and Altman plot was used to illustrate the difference and mean score of the testing results.[23] The part about value assessment to health care voucher was used to test the reliability of the questionnaires among older people and optometrists. A satisfactory percentage agreement (≥ 70%) was noted in all items. The difference of test-retest results was within 95% of agreement across the mean result.

The study was approved by the Human Subjects Ethics Sub-committee of The Hong Kong Polytechnic University and conducted in accordance with the tenets of the Declaration of Helsinki. An information sheet was given to each subject and written consent obtained before data gathering was initiated.

### Patient and Public involvement

No patient involved.

## STATISTICAL METHODS

Descriptive statistics used included frequencies and percentages summarizing the demographic information of subjects, pattern of optometry services and voucher utilization and optometrist’s perception of the value of voucher scheme. The association of the interval of last eye check between those who used and did not use the EHV for optometry services was evaluated by the Chi-square test. The Mann Whitney U test was used to test the differences in access barriers between those who had and did not have regular ocular health assessment. Data was analyzed using IBM SPSS statistics version 25 software (SPSS Inc., Chicago, Illinois).

Thematic analysis was used to analyze the results of the face-to-face interview. The six phrases of thematic analysis included (1) Familiarization with the data; (2) Generating initial codes; (3) Searching for themes; (4) Reviewing themes; (5) Defining and naming themes; and (6) Producing the report.[24] The themes and sub-themes were identified from recurring topics in the transcripts.

## RESULTS

Between November 2017 and August 2018, 1176 subjects were recruited in the first part of study. The response rate was 100%. There were 20 incomplete questionnaires collected, resulting in 1156 (98.3%) valid questionnaires for analysis. Among the recruited subjects, 650 (56.2%) were female, aged 70 to 74 years (33.8%) and attained either primary (42.8%) or secondary (36.2%) education level. Around 60% of the participants had 1 to 2 medical conditions, including diabetes, hypertension, and hyperlipidemia, among others. The demographic information is shown in Table 1. Twenty subjects participated in the face-to-face interview.

**Table 1.** Demographics of the subject population (n = 1156)

| Demographics | Frequency | % |
| --- | --- | --- |
| Gender |  |  |
| Male | 506 | 43.8 |
| Female | 650 | 56.2 |
| Age |  |  |
| 65-69 | 294 | 25.4 |
| 70-74 | 391 | 33.8 |
| 75-79 | 288 | 24.9 |
| 80-84 | 113 | 9.8 |
| 85 or above | 70 | 6.1 |
| Education |  |  |
| No formal schooling | 168 | 14.5 |
| Primary | 495 | 42.8 |
| Secondary | 418 | 36.2 |
| Post-secondary | 53 | 4.6 |
| Degree or above | 22 | 1.9 |
| No. of medical condition(s) |  |  |
| 0 | 406 | 35.1 |
| 1 | 434 | 37.5 |
| 2 | 231 | 20.0 |
| 3 | 78 | 6.7 |
| More than 3 | 7 | 0.6 |

In the second part of this study, 389 optometrists completed the questionnaire, representing 38% of eligible optometrists and 58% of those who had enrolled in the Scheme during the study period. Most of the respondents were in the age group of 30-39 years (39.3%) (Table 2). Among the 286 optometrists who had enrolled in the Scheme, 47.6% indicated that they worked in optical shop settings and 43.4% worked in optometry clinic settings. For the 103 optometrists who were not enrolled in the Scheme, 44.7% indicated they were working in public hospitals or clinics where EHV was not applicable (Table 3).

**Table 2.** Demographics of optometrists participating in this study (n= 389)

| Demographics | Frequency | % |
| --- | --- | --- |
| Gender |  |  |
| Male | 198 | 51.0 |
| Female | 191 | 49.0 |
| Age |  |  |
| 20-29 | 128 | 32.9 |
| 30-39 | 153 | 39.3 |
| 40-49 | 77 | 19.8 |
| 50-59 | 26 | 6.7 |
| 60 or above | 5 | 1.3 |

**Table 3.** Type of practice settings (n = 389)

| Type of practice | Enrolled in EHV scheme | Not enrolled in EHV scheme |
| --- | --- | --- |
| Hospital Authority/<br>Department of Health | 2 (0.7%) | 46 (44.7) |
| Industry | 0 (0%) | 2 (1.9%) |
| Not practicing | 1 (0.3%) | 2 (1.9%) |
| Ophthalmology clinic | 10 (3.5%) | 12 (11.7%) |
| Optometry clinic | 124 (43.4%) | 10 (9.7%) |
| Private hospital | 1 (0.3%) | 10 (9.7%) |
| Optical shop | 136 (47.6%) | 18 (17.5%) |
| Teaching/research | 12 (4.2%) | 3 (2.9%) |
| Total | 286 (100.0%) | 103 (100%) |

### Pattern of eye examinations

About 1 in 3 subjects indicated that they did not have regular eye examinations (38.4%). Some subjects had their eyes examined once every year (22.8%) or every two years (15.6%). Around half of them had their last examination within the past 2 years (53.7%). However, 22% of subjects reported that they have not had their eyes examined before.

The most common reason of seeking for an eye examination was “having trouble seeing” (33.4%), followed by the need for a new prescription for spectacles (23.8%), referral by other health care providers (9.1%) and eye discomfort (4.9%) or other eye problems (2.8%). Public clinics or hospitals (37.3%) and optical shops (25%) were most commonly used settings by subjects for their eye examinations. Fewer subjects went to private clinics or private hospitals (21.6%) and non-government organizations (15.1%).

For those who have not had an eye examination before, around half of them considered their vision acceptable. The remaining were concerned about the relative costs of the service or transportation fees (21.8%) and 18.3% of them were still waiting for their scheduled appointment. Some did not know where to get an eye examination (8.9%).

### Access barriers to optometry services utilization

Our subjects were asked to indicate their access barriers to optometry services using a 5-point Likert scale. Uncertainty about consultation fees and cost of spectacles were the highest ranking (median = 4.0). The access barriers were compared between those subjects who used and who had not used optometry services (Mann-Whitney U test). All barriers including consultation fees, cost of spectacles, location of service providers, appointment waiting time, transportation, accompanying person and information about the availability of service were significantly different between the two groups (Table 4, 5).

**Table 4.**
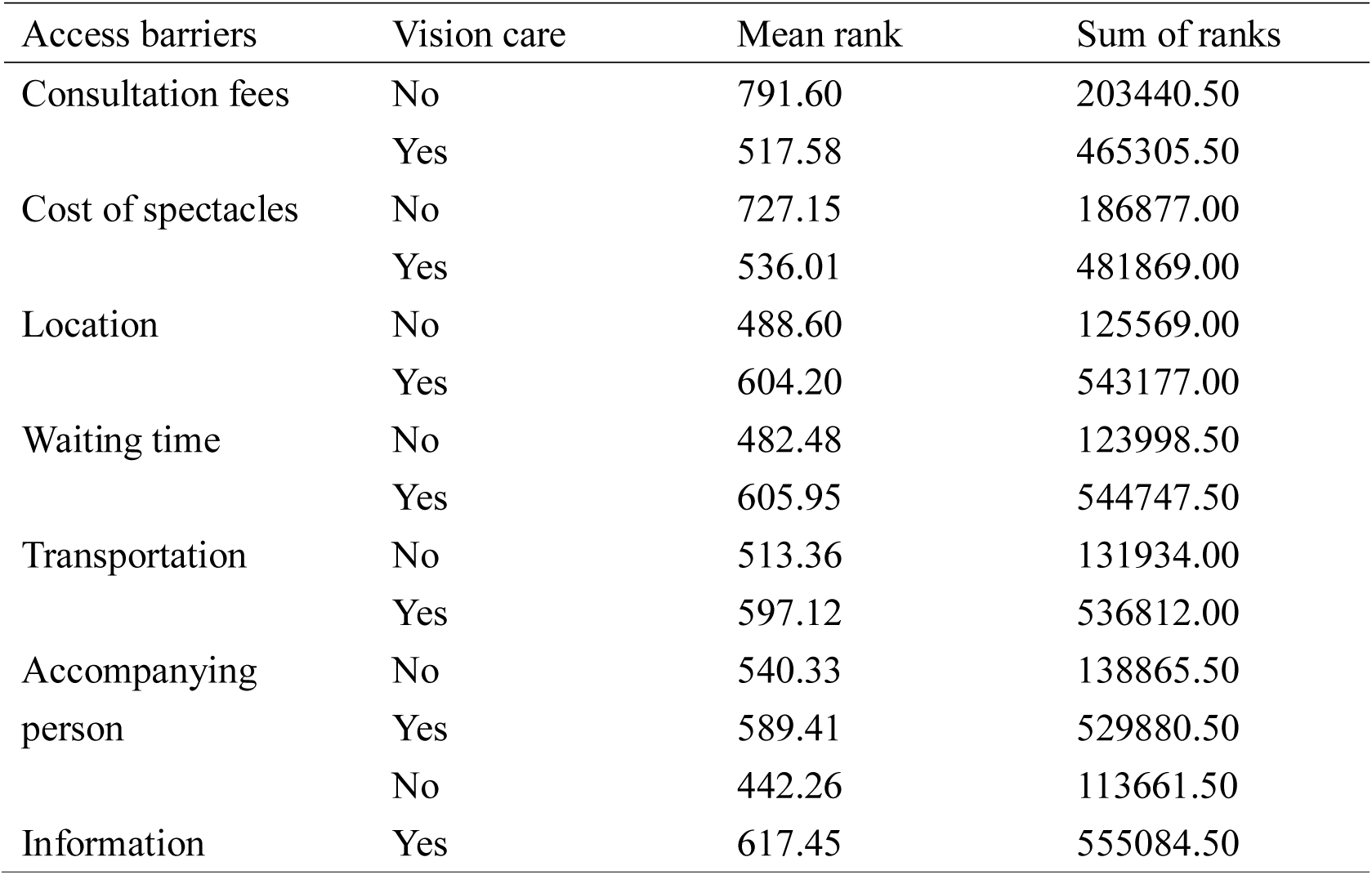
Mean rank and sum of ranks of access barriers

| Access barriers | Vision care | Mean rank | Sum of ranks |
| --- | --- | --- | --- |
| Consultation fees | No | 791.60 | 203440.50 |
|  | Yes | 517.58 | 465305.50 |
| Cost of spectacles | No | 727.15 | 186877.00 |
|  | Yes | 536.01 | 481869.00 |
| Location | No | 488.60 | 125569.00 |
|  | Yes | 604.20 | 543177.00 |
| Waiting time | No | 482.48 | 123998.50 |
|  | Yes | 605.95 | 544747.50 |
| Transportation | No | 513.36 | 131934.00 |
|  | Yes | 597.12 | 536812.00 |
| Accompanying person | No | 540.33 | 138865.50 |
|  | Yes | 589.41 | 529880.50 |
| Information | No | 442.26 | 113661.50 |
|  | Yes | 617.45 | 555084.50 |

**Table 5.** Mann-Whitney U test results

|  | Consultation<br>fee | Spectacle<br>fee | Location | Waiting<br>time | Transport | Accompany | Information |
| --- | --- | --- | --- | --- | --- | --- | --- |
| U | 60755.5 | 77319.0 | 92416.0 | 90845.5 | 98781.0 | 105712.5 | 80508.5 |
| Z | -12.097 | -8.424 | -5.020 | -5.353 | -3.629 | -2.183 | -7.671 |
| P | <0.005 | <0.005 | <0.005 | <0.005 | <0.005 | 0.029 | <0.005 |

From the face-to-face interview, insufficient access to information about services was revealed as a barrier to access of optometry services. Some subjects mentioned that they did not know the fees charged in the private sector and thought it was generally unaffordable. One interviewee aged 68 years said, ‘I am not sure if the consultation fees in the private sector will be higher if I have more than one eye disease.’ Some subjects preferred to stay with the public sector because they were not sure which private providers offer comprehensive services or which ones to trust. This echoes the findings from the questionnaire results that consultation fees and information on the availability of service were access barriers for optometry services.

### Using the EHV for optometry services

The questionnaire results showed that almost all subjects (97.5%) knew about the EHV scheme. However, 20.3% of subjects did not use the voucher for any health care service. The subjects were asked to rank their preferences of using the voucher on services provided by health care professionals. Medical services had the highest ranking (86.9%), followed by dental services (41.0%) and optometry services (39.0%). Some subjects did not show specific preferences (7.5%).

The time of last eye examination was found to be associated with the usage of voucher for optometry services (Chi square = 27.889, p < 0.001 with df = 2). Those who had used the EHV on optometry services before had eye checks more regularly than those who did not. Among those who had an eye examination before, 80.4% of EHV users of optometry services had their last eye examination within 2 years, while it was only 64.1% in non-EHV users of optometry services.

Among those who had used vouchers for optometry services previously, 10.7% were used for consultation alone. The majority of the subjects also used the vouchers for prescription spectacles (87.4%). Some used the voucher for eye drops (11.2%), sunglasses (6.2%) and visual rehabilitation aids (0.3%).

Subjects were asked to comment on the effectiveness of the EHV scheme on a 5-point Likert scale. Around half of the subjects who had heard about the Scheme agreed it could facilitate them to have regular eye examinations, obtain prescription aids when necessary and have a better understanding of the condition of their eyes (4 points or above).

### Barriers to EHV use

Although optometry service was ranked top 3 preferred health care services under the EHV scheme, only 353 (39.3%) subjects in this study used their vouchers on services provided by optometrists. Most subjects who had not used the voucher for vision care service mentioned that they thought the voucher value was insufficient for the consultation fee or costs of prescription spectacles (21.7%). Some subjects used the public service where voucher could not be used (17.1%). Some were satisfied with their eye condition and felt they did not need an eye check (14.3%). Others preferred to use the vouchers on other health care services (11.7%) or their optometrists were not enrolled on the EHV scheme (10.8%). The face-to-face interview showed a similar result that subjects preferred to spend their vouchers on curative services, including acute diseases and existing chronic disease management. Subjects suggested that the dollar value of the vouchers should be increased to meet their health needs.

### Value of the EHV scheme-perspective from optometrists

Almost all optometrists (97.5%) agreed that the EHV scheme was beneficial to the optometry profession. Over 80% of the optometrists agreed the scheme had improved awareness of eye conditions and could facilitate early detection of sight-threatening eye diseases. Moreover, 95.5% of the optometrists agreed that the scheme enabled older people to obtain prescription spectacles where necessary.

To improve the usage and effectiveness of the voucher, optometrists suggested a closer monitoring of voucher usage to avoid abuse, more public education on voucher use and increasing the voucher value.

## DISCUSSION

### Optometry services among top 3 preferred health services

An optometry service was ranked top three preferred services when using the EHV. This is within expectation as it has been shown in a recent US-based survey of adults that eye health was considered by most people to be an integral part of general health and for around half the people surveyed, loss of sight was the worst possible health outcome.[25]

### The EHV enabled more elderly people to access optometry services

From our study, older people who had used the voucher before had more regular eye examination than those who had not. Timely management of eye abnormalities could be facilitated with regular eye examinations. However, our findings showed that only about 40% of subjects had used the voucher on the optometry services, meaning that the majority 60% chose not to use their vouchers on optometry services. Nevertheless, figures provided by the Hong Kong SAR government on EHV utilization show that in the “HK$ 500 or below” category of claims for optometry services (the low dollar value is likely to reflect just the consultation fees), there were 3577 transactions in 2014 and 26883 transactions in 2018 [19]. This suggests that increasingly more users are having their eyes examined under the EHV scheme. In our survey, 10.7% of our subjects who used the EHV for optometry services used the voucher for consultation fees alone. The vast majority also had optical aids (usually spectacles) dispensed.

Lowering the financial barrier to accessing vision-related services clearly encouraged more eligible users of the EHV to use optometry services. In Hong Kong, optometry services can be accessed directly without referral, in contrast to some of the other health care providers (for example, physiotherapists) of the EHV scheme, where a referral from a medical practitioner is required. Those eligible EHV users who are using public hospitals and specialist clinics have the option to seek vision-related services in the private sector, decreasing delay in service access.

### Other access barriers still preventing access to optometry services

The 60% of subjects who did not use the EHV on optometry services identified many other access barriers which discouraged or prevented them from using the services. Uncertainty about consultation fees, cost of spectacles, services available and location of enrolled service providers are significant barriers to potential service users. Our results from the questionnaire survey and focus group interview clearly show a need for price transparency in order to remove a major access barrier to optometry services. In the Hong Kong setting, the usual practice is for the various consultation and dispensing charges to be totaled up and presented to the service user at the very end. Uncertainty about the cost of health care service is a known barrier to service utilization in the older population.[26] Better price transparency could improve healthcare service utilization and users would have more confidence to seek service.[27] To improve price transparency in optometry services, consultation and other service fees, and the range of charges for some common services and products (such as single vision lens and progressive lens) could be published on the websites of individual optometry practices and optical shops, as well as professional associations.

Interestingly, these barriers are within the capability of the service providers, collectively or individually, to lower or minimize. The fact that it has not been dealt with yet suggests the current situation favours the supply side. There are over 1.2 million eligible EHV users and 780 optometrists enrolled in 2019.[20, 28]

### Annual EHV dollar allocation should be increased

The results from the questionnaire and face-to-face interview showed that our subjects considered the current voucher amount was insufficient and they preferred to use the voucher on curative services. This resonates with the results of local studies on the health care voucher.[14, 15] It is not surprising that users would hold this view. Most elderly people have some combinations of chronic diseases, and in addition, age-related dental and vision needs. The annual voucher allocation seemed small in relation the potential costs associated with managing these conditions in the private sector. To promote a greater use of preventive services would require further tuning of the EHV scheme such as injecting a higher amount to the annual EHV allocation or appropriating parts of the EHV allocation to targeted preventive services, amongst other possibilities.

### Service providers

The majority of the eligible optometrists enrolled as service providers in the EHV scheme. Those who did not enroll were working in public clinics or hospitals where the voucher could not be used. Our sample of optometrists agreed the voucher scheme was useful to raise the awareness of vision care and facilitate vision correction by spectacles among the older population. While the financial aspects of the EHV are no doubt attractive to optometry services providers, they also have a critical role to play to lower other access barriers for EHV users such as providing price transparency and public education on the threats of poor vision on daily living activities and the benefits of early screening for potentially sight-threatening conditions.

The EHV scheme provided an opportunity for the government to raise the quality of optometry services. The scheme only enrolled those optometrists qualified for full scope practice. Enrolled optometrists, as part of the agreement with the government, have to provide optometry services in a professional manner, a re-enforcement of the code of practice of the optometrist statutory registration body. The EHV scheme office receives complaints from service users and takes appropriate action. The scheme office also conducts random audits, which although mostly financial in nature, nevertheless potentially promotes discipline and habit-forming compliance to regulations. As the funding agency for the EHV, the government could reasonably expect service quality improvements and set desired targets to reflect these quality improvements.

### Recent policy adjustments in relation to optometry services

Statistics on voucher use in 2018 show that the median voucher amount per claim by optometrist was the highest among all the eligible professions (HK$ 1,951), while the amount from medical practitioners and dentists was HK$ 330 and HK$ 640 respectively.[20] Since the majority of the older people use the voucher on spectacles, the annual entitlement of HK$ 2,000 would be used up easily. Against the underpinning philosophy of the EHV scheme which espoused the freedom for users to choose primary care services that suited their needs offered by approved practitioners[29], the government took the view that over-concentration of EHV spending on optometry services, particularly for prescription optical aids, was undesirable. It implemented a voucher spending ceiling of $ HK2,000 every two years for optometry services in 2019. The accumulation ceiling was raised from $ 5,000 to HK$ 8,000 in 2019. This achieved the desired effect and the median claim for optometry services in 2019 dropped to HK$ 1,750. More significantly, the total sum claimed for optometry services dropped from HK$ 760 million in 2018 to $ 432 million in 2019.[20]

Setting a ceiling on spending would clearly shape spending behavior. However, it may also discourage people from using optometry services as they may need to pay part of the fees out of pocket. Further evaluation should be conducted by the government and the optometric profession to monitor the change of service utilization.

### Strengths and limitations of this study

The strength of this study is the high representativeness of community dwelling older population in Hong Kong. Our subjects were recruited by cluster sampling covering community centers in all the 18 districts of Hong Kong. The number of participants recruited was in proportion to the total number of older population in each district. However, we recognized that the characteristics and the pattern of vision care utilization of the older people recruited from community centers could be different from those staying at home. Further population-based research with a larger sample size could provide a more accurate picture of the health service seeking behavior using the EHV. The mixed method approach of this study provided a more in-depth investigation of the population. Data collection by questionnaire is efficient and draws conclusion from a large group. The qualitative part by face-to-face interview enriches the information explores individual’s experience which is not possible to list in a questionnaire.

## CONCLUSION

The EHV scheme promoted greater access and reduced the financial barrier to optometry services. Through this scheme, every eligible resident of Hong Kong aged 65 years or above has the financial means to choose to access optometry services, including provision of basic prescription optical aids. However, other non-financial barriers remain and service providers could play a more active role to reduce these barriers by providing more information about available optometry services, improving transparency of consultation fees and cost of spectacles.

## Data Availability

The data in the study was processed and transferred to the information in the manuscript

## Acknowledgement

Yap M is supported by the KB Woo family endowed professorship.

## Author statement

Sum R designed the protocol, analyzed the data and wrote the manuscript. Yap M contributed to the data interpretation and revised the manuscript. Both authors approved the final version to be published.

